# MVA-BN Vaccine Effectiveness: A Systematic Review of Real-World Evidence in Outbreak Settings

**DOI:** 10.1101/2024.04.25.24306078

**Authors:** Lauren M.K. Mason, Estefania Betancur, Margarita Riera-Montes, Florian Lienert, Suzanne Scheele

## Abstract

**Background:** Mpox is a disease endemic to Central and West Africa. It caused outbreaks in non-endemic countries, mainly in 2022. The endemic Democratic Republic of Congo is currently experiencing its largest outbreak yet. The vaccine Modified Vaccinia Ankara-Bavarian Nordic (MVA-BN) is approved for active immunization against mpox and smallpox. Since the outbreak in 2022, real-world studies have assessed MVA-BN’s vaccine effectiveness (VE) against mpox, and this systematic literature review aims to summarize the most current evidence.

**Methods:** Medline (via PubMed), Embase, and LILACS were searched, as well as grey literature sources and publications’ bibliographies to identify observational studies published between 1/Jan/2022 and 28/Feb/2024 that estimate the VE of MVA-BN against symptomatic mpox or provide risk measures that allow calculation of these VE estimates. Data were presented descriptively in tables and text; the methodological quality of included records was assessed using an informal qualitative approach.

**Results:** The literature search identified a total of 16 records that fit the inclusion criteria. The studies took place in high-income countries and were heterogenous in design, setting, and definition of at-risk populations. MVA-BN VE estimates against symptomatic mpox infection ≥14 days post-vaccination were assessed. Where the study population was exclusively or primarily those receiving pre-exposure prophylactic vaccination, the adjusted VE estimates ranged from 35% to 86% (n=8 studies) for one dose and from 66% to 90% (n=5) for two doses. Where only post-exposure prophylactic vaccination was assessed, adjusted VE estimates were reported for one dose only at 78% and 89% (n=2). Additionally, MVA-BN reduced the risk of mpox-related hospitalization in one study and the severity of mpox clinical manifestations in two studies.

**Conclusions:** Despite heterogeneity in study design, setting, and at-risk populations, the reported VE estimates against symptomatic mpox infection for one or two doses of MVA-BN support deployment of MVA-BN for mpox outbreak control.

## Introduction

Mpox, formerly called monkeypox and first described in humans in the Democratic Republic of Congo (DRC) in 1970 [1], is an illness that was initially zoonotic and is caused by the monkeypox virus (MPXV). This DNA virus belongs to the *Orthopoxvirus* genus, which also includes the smallpox-causing variola virus [2]. The clinical presentation of mpox includes a distinctive, extensive rash, fever, headache, cough, asthenia, and lymphadenopathy. Additionally, complications such as coalescence of skin ulcers, bacterial skin infections, bronchopneumonia, and sepsis may occur [3]. MPXV is transmitted from animals to humans and between humans through contact with bodily fluids, lesions on the skin or mucosae, respiratory droplets, and contaminated objects [4]. A definitive animal reservoir host has not been identified [1].

Before the unprecedented global spread of the disease outside previously endemic countries in 2022, reports of mpox were almost exclusively limited to Central and West African countries [4]. An observed increase in cases in these countries before 2018 was hypothesized to be due to the cessation of routine smallpox vaccination leading to waning *Orthopoxvirus* immunity [1].

Starting in May 2022, a significant surge in mpox cases occurred in numerous non-endemic countries [5]. As of February 2024, over 94,000 confirmed mpox cases have been documented in 117 countries [6]. While previous occurrences of mpox in non-endemic regions were associated with international travel and importation of infected animals from West Africa [7], the 2022 outbreak was primarily characterized by human-to-human transmission of MPXV [8]. This outbreak mainly occurred among, but was not limited to, men who have sex with men (MSM) and in several high-income countries [9], which had not been reported previously. Other differences included a different clade type, lower mortality, and suggested novel epidemiological and clinical characteristics [9]. The outbreak response strategy consisted mainly of behavior change and vaccination of high-risk populations [2].

The DRC is currently experiencing its largest ever recorded mpox outbreak, with more than 12,000 suspected cases and 500 deaths reported since the start of 2023 [10]. The outbreak is spreading to almost all provinces, with different transmission patterns and affected populations. Some areas are seeing large-scale transmission involving a larger proportion of children, potentially due to contact with rodents. The province of Equateur, with a population of almost 6 million, is experiencing large, concomitant mpox and measles outbreaks, with a potential increase in the risk of complications and a larger reported mortality rate, as well as the risk of misdiagnosis of either disease. There is also a large outbreak of primarily sexually transmitted mpox infections among sex workers and adults in the mining city of Kamituga in South Kivu. The city’s highly mobile population creates the risk for a multi-country outbreak as the area borders Rwanda, Burundi, and Tanzania [11]. Global outbreak response teams from multiple countries are active in the DRC, and mpox vaccination may soon become a tool in controlling the current crisis.

Modified Vaccinia Ankara-Bavarian Nordic (MVA-BN; trade names Jynneos/Imvanex/Imvamune) is the smallpox and mpox vaccine that has most broadly been used in response to the global mpox outbreak. MVA-BN cannot replicate in human cells and, unlike replicating smallpox vaccines, has a favorable safety profile for individuals with atopic dermatitis and immunodeficiency [12].

The MVA-BN vaccine can be administered prophylactically either before or after exposure to confirmed mpox cases. Pre-exposure prophylactic vaccination (PrEP) aims at protecting those at risk of mpox infections prior to having had contact with a confirmed case. Currently, the US Centers for Disease Control and Prevention (CDC) recommends PrEP for the at-risk groups: those at risk of occupational exposure, gay, bisexual, and other MSM (GBMSM) who have been diagnosed with certain sexually transmitted diseases and have multiple sexual partners, individuals engaging in sexual activities in areas with a high prevalence of mpox, and those with sexual partners meeting these criteria, as well as people with human immunodeficiency virus (HIV) infection or other causes of immunosuppression who have had recent or anticipate potential mpox exposure [13]. Post-exposure prophylactic vaccination (PEP) aims to prevent mpox or reduce its severity in individuals who have had contact with confirmed cases and can be administered up to 14 days post-exposure [13].

The regulatory authorities in the US, Canada, UK, and the EU approved MVA-BN in a two-dose schedule ≥28 days apart for active immunization against smallpox and mpox based on pre-clinical efficacy data from animal studies and clinical immunogenicity and safety studies [14–17]. The efficacy of MVA-BN against mpox has not been demonstrated in randomized controlled clinical trials; however, since the 2022 outbreak started, several real-world effectiveness studies have been conducted. This systematic literature review (SLR) aims to identify and summarize the most current real-world evidence (RWE) on the MVA-BN vaccine effectiveness (VE) against symptomatic mpox infection, mpox-related hospitalization, and severity of clinical mpox manifestations.

## Methods

### Registration and protocol

This SLR was registered prospectively in PROSPERO (https://www.crd.york.ac.uk/prospero/; number CRD42023441204), where key features from the protocol can be retrieved. The literature search was initially performed in 2023 and updated on February 28, 2024, to include the latest relevant literature. The focus of this manuscript is on the summary of RWE of the VE of MVA-BN against mpox.

The protocol and manuscript for this SLR were developed in line with recommendations from the Preferred Reporting Items for Systematic Reviews and Meta-Analyses (PRISMA) statement and PRISMA extension for protocols (PRISMA-P) [18, 19].

### Search strategies and eligibility criteria

The databases Medline (via PubMed), Embase, and LILACS were searched using database-specific keywords and medical subject headings (MeSH terms) related to mpox and vaccine effectiveness/vaccination. The detailed search strategies are available in **Supplementary Methods**. We included peer-reviewed publications reporting on observational studies in humans on the VE of MVA-BN, published between January 1, 2022, and February 28, 2024 (date of last search) in English, Spanish, or French. We excluded publications that did not meet the inclusion criteria or where methods and sources for data collection and/or analysis were not clearly defined. Records reporting on *in vitro* studies, modeling studies, case reports/series, and (non-)randomized clinical trials were excluded. SLRs and meta-analyses (MAs) were retained through the search and first selection stage to allow screening through the bibliographies for identification of additional relevant publications, but SLRs and MAs were not retained for data extraction. The full list of eligibility criteria is available in **Supplementary Methods**.

To identify additional relevant publications, grey literature sources and pre-print databases were searched on March 5, 2024 (date of last search) using source-specific search terms and limits as detailed in **Supplementary Methods**.

### Record selection

After deduplicating identified records, two independent researchers (EB, LM) screened the titles and abstracts of all records in duplicate. Results were compared and discrepancies were discussed; in case of doubt, the record was included for full-text review. Records for which insufficient information was available in the title and abstract were also included for full-text review. All full texts were reviewed by one researcher (EB), and full texts for 10% of the records were screened in duplicate (EB, LM). The results were compared and discrepancies were discussed; any doubts outside the 10% were also discussed. When multiple articles reported on the same dataset and the same outcome, only the most recent and complete available version was included. To identify additional relevant publications, bibliographies of selected papers (e.g., key studies, SLRs, and MAs) were manually searched.

### Data collection and synthesis

One reviewer (EB) extracted all relevant data for each retained record in a pre-defined template in Microsoft Excel. The main outcomes of interest were crude and/or adjusted VE estimates against symptomatic mpox infection, mpox-related hospitalization, and severity of mpox clinical manifestations with crude/adjusted confidence intervals (CIs). Data related to these VE estimates, such as VE calculation methods, subpopulations, stratification groups, incidence rates, cumulative hazard rates, risk association measures (odds ratios [ORs], risk ratios [RRs], hazard ratios [HRs], incidence rate ratios [IRR], hazard rate ratios [HRRs]), and confounders were also extracted. In case no crude or adjusted VE estimates were reported in the publication, we calculated the VE and 95% CIs from extracted risk association measures using the following formula:

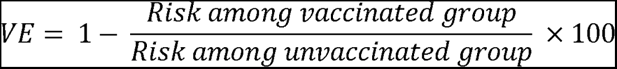

No other calculations or assumptions were made in case of other missing data. Variables related to study characteristics (e.g., study design, vaccination strategy), population characteristics (e.g., sample definitions, comorbidities), study methods (e.g., case and control definitions), additional information on statistical analyses (e.g., results of the sensitivity analysis), and data for assessment of the study’s methodological quality (e.g., strengths and limitations as mentioned by the authors) were also extracted.

Study characteristics and the collected VE estimates were presented descriptively in tables and text. VE estimates were reported stratified by type of vaccination strategy: PrEP or PEP. For the former, we reported studies including exclusively PrEP recipients together with studies designed primarily to evaluate PrEP, but that may have inadvertently included some PEP subjects (designated ‘PrEP ± PEP’). Due to the heterogeneity of the data, no statistical synthesis of the extracted data was performed.

### Risk of bias assessment

The methodological quality of all records included in the final review was assessed by three researchers (EB, LM, MRM) using an informal qualitative approach based on the expertise of reviewers and published commentaries, as well as the limitations described by the authors themselves. Potential biases of the studies were identified and how these might impact the results and conclusions of the studies was considered and discussed.

## Results

### Study selection and characteristics

The literature search identified 904 unique records from databases, of which 106 were retained for full-text review and eight were included in the final review. Nine additional records were identified from other sources, of which eight were included in the final review. Hence, a total of 16 records on 16 unique studies were identified that reported on the VE of MVA-BN against symptomatic mpox infection or provided risk measures that allow calculation of VE estimates (**Figure 1**).

**Figure 1.**
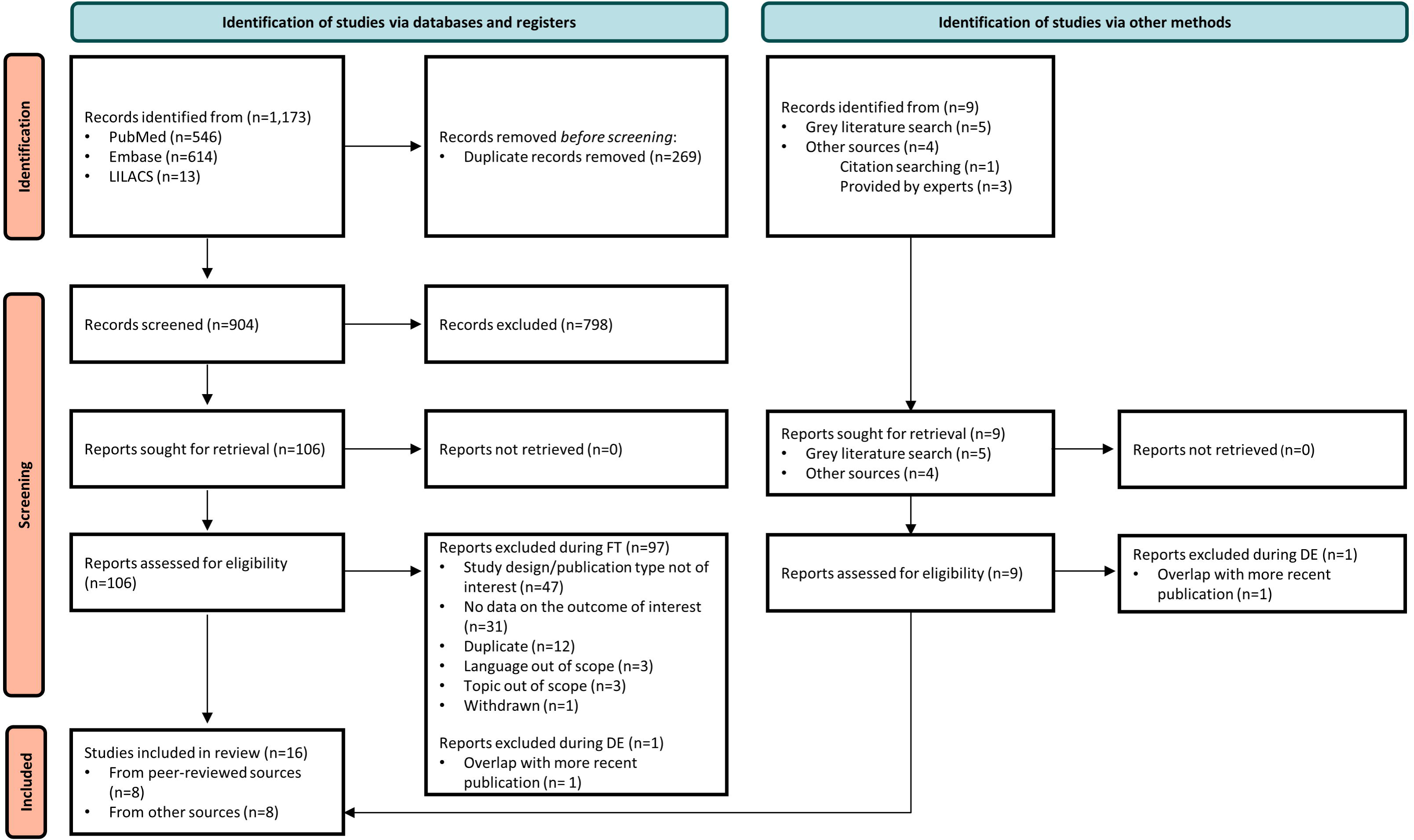
PRISMA flowchart. DE: data extraction; FT: full-text review

The main characteristics of the included studies are summarized in **Table 1**. The studies took place in six countries, with the majority conducted in the United States (n=7) [20–26], followed by Canada (n=3) [27–29], Spain (n=2) [30, 31], the United Kingdom (n=2) [32, 33], and one each in Israel [34] and the Netherlands [35]. The studies used various designs to assess effectiveness of MVA-BN, with a retrospective cohort design being used most frequently (n=6) [23, 24, 26, 27, 30, 34], followed by a case-coverage design (n=4) [22, 32, 33, 35], a case-control design (n=4; incl. one test-negative case-control design [28]) [20, 21, 25, 28], and a prospective cohort design (n=2) [29, 31]. The studies used linked data from public health surveillance systems [20, 22–30, 32, 33], from public health institutions [23, 31, 35], and/or from healthcare systems [21, 34]. The number of mpox cases included for calculation of the VE against symptomatic mpox infection ranged from 137 to 11,320. Control groups were defined in various ways across study designs and included MSM, incident HIV cases, males with sexually transmitted infections (STIs), symptomatic individuals who tested negative for mpox, and individuals in contact with mpox cases. The study period covered (part of) 2022 in 11 studies [21–25, 28, 30–32, 34, 35], (part of) 2022 and part of 2023 in four studies [20, 26, 27, 29], and the whole of 2023 in one study [33]. Fourteen studies focused on the use of MVA-BN as PrEP, of which six included PrEP recipients only [21, 28–30, 32, 33] and eight may have included a proportion of PEP recipients due to the available data not allowing exclusion of all PEP recipients (PrEP ± PEP) [20, 22, 23, 25–27, 34, 35]; the remaining two studies focused on the use of MVA-BN as PEP [24, 31]. The effectiveness of MVA-BN against mpox was evaluated using ORs (n=10) [20, 21, 24–28, 32, 33, 35], HRs (n=4) [23, 29, 31, 34], RRs (n=1) [30], or IRRs (n=1) [22] (**Table 1**).

**Table 1.**
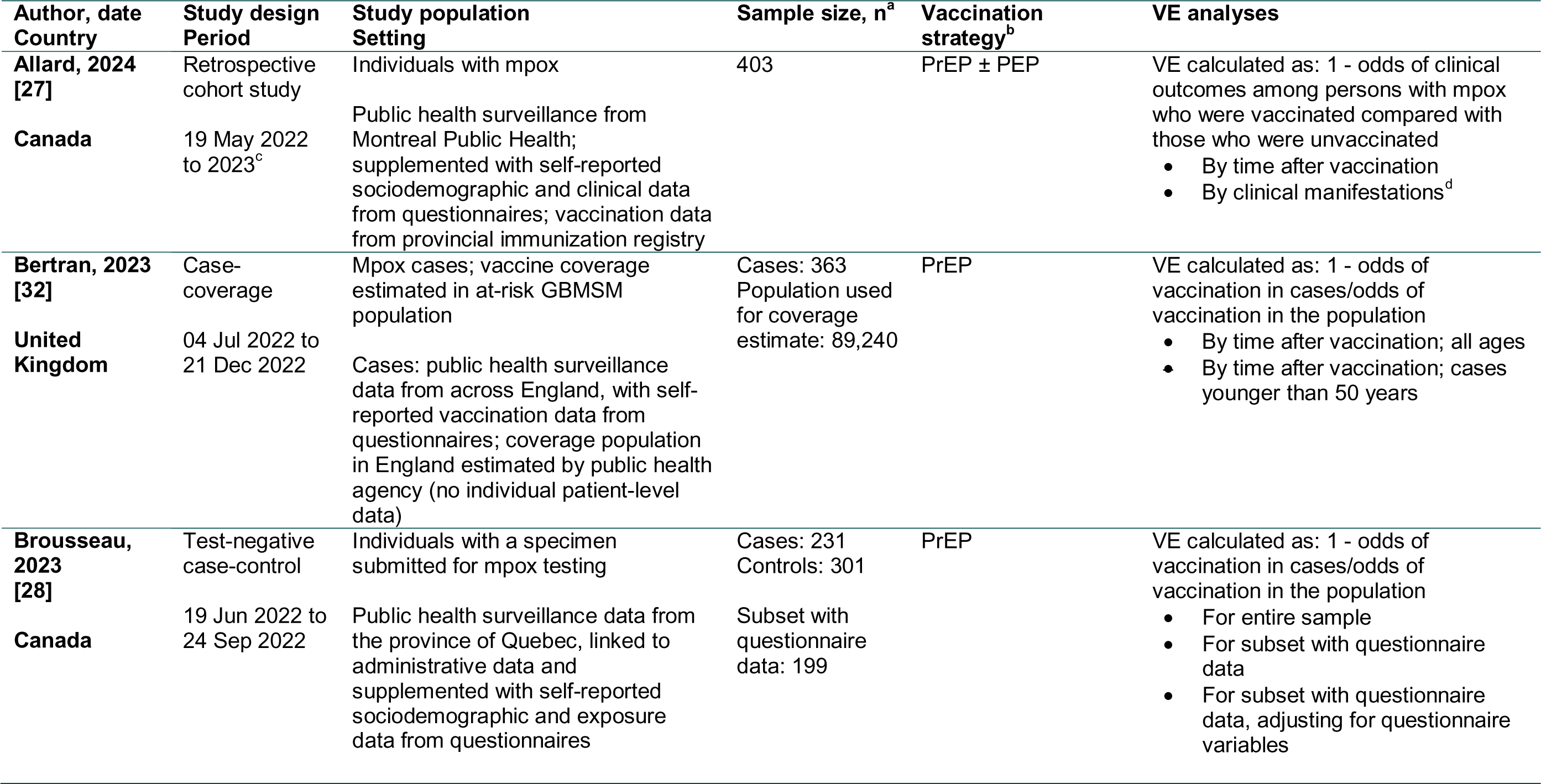
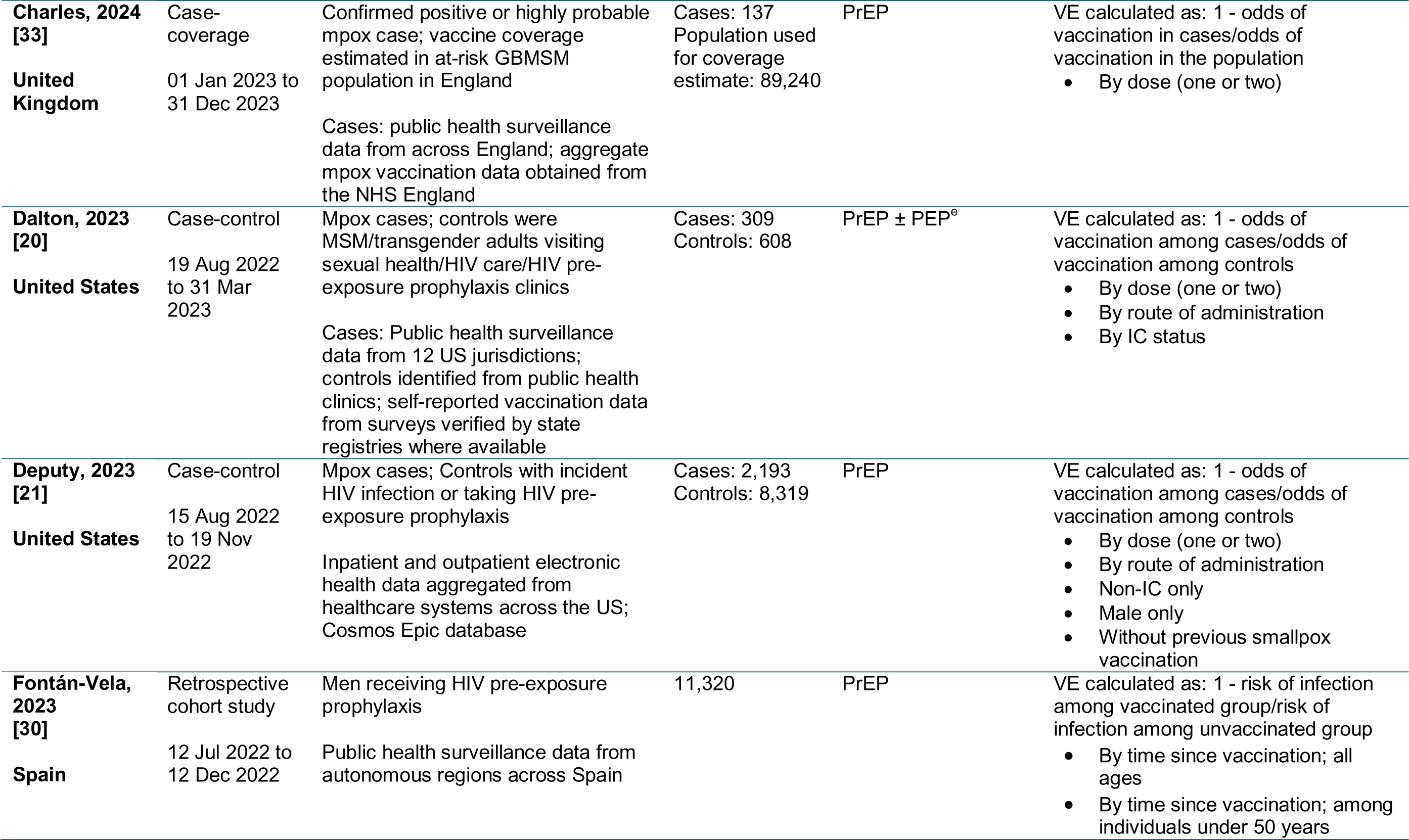
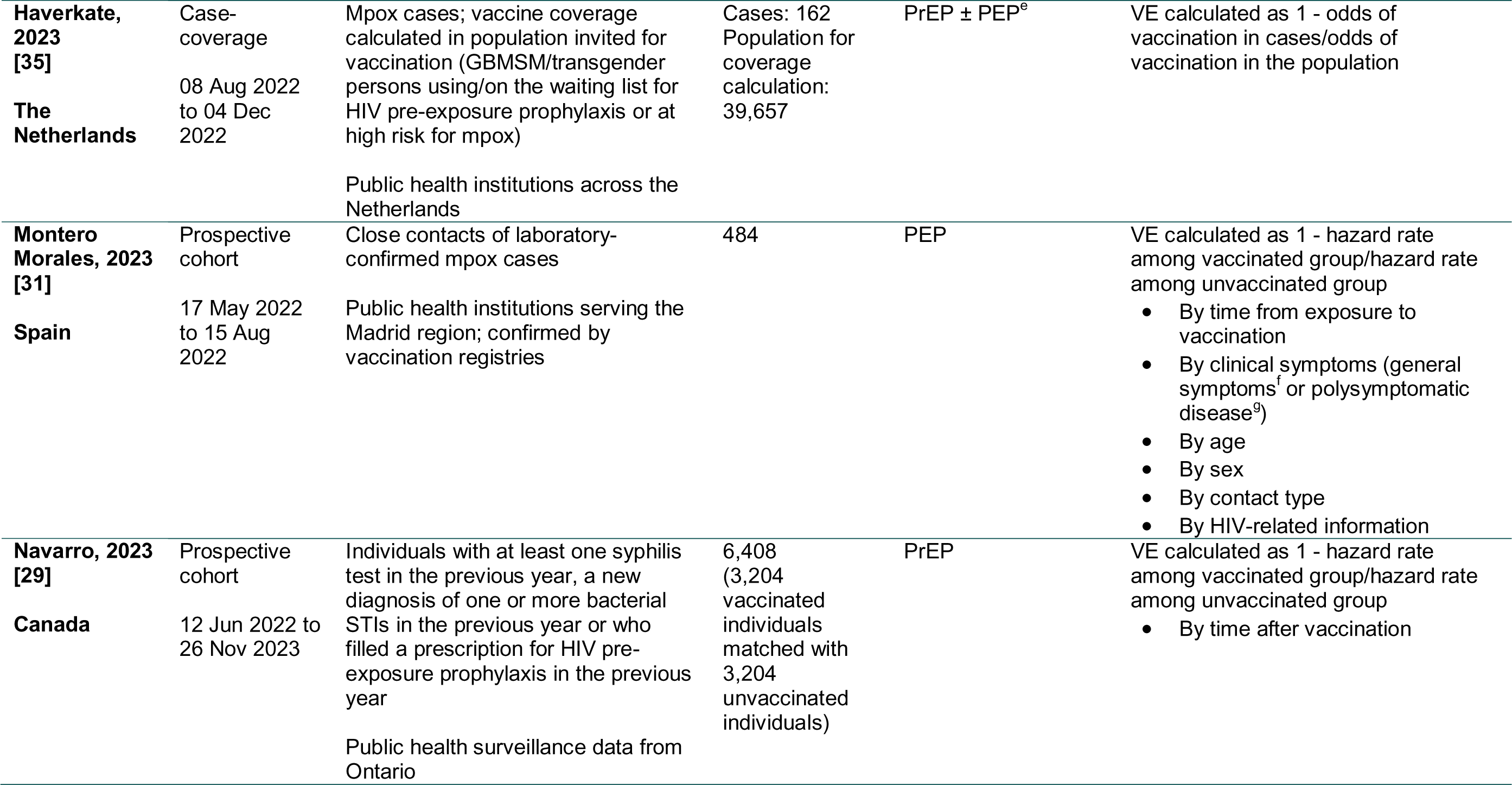
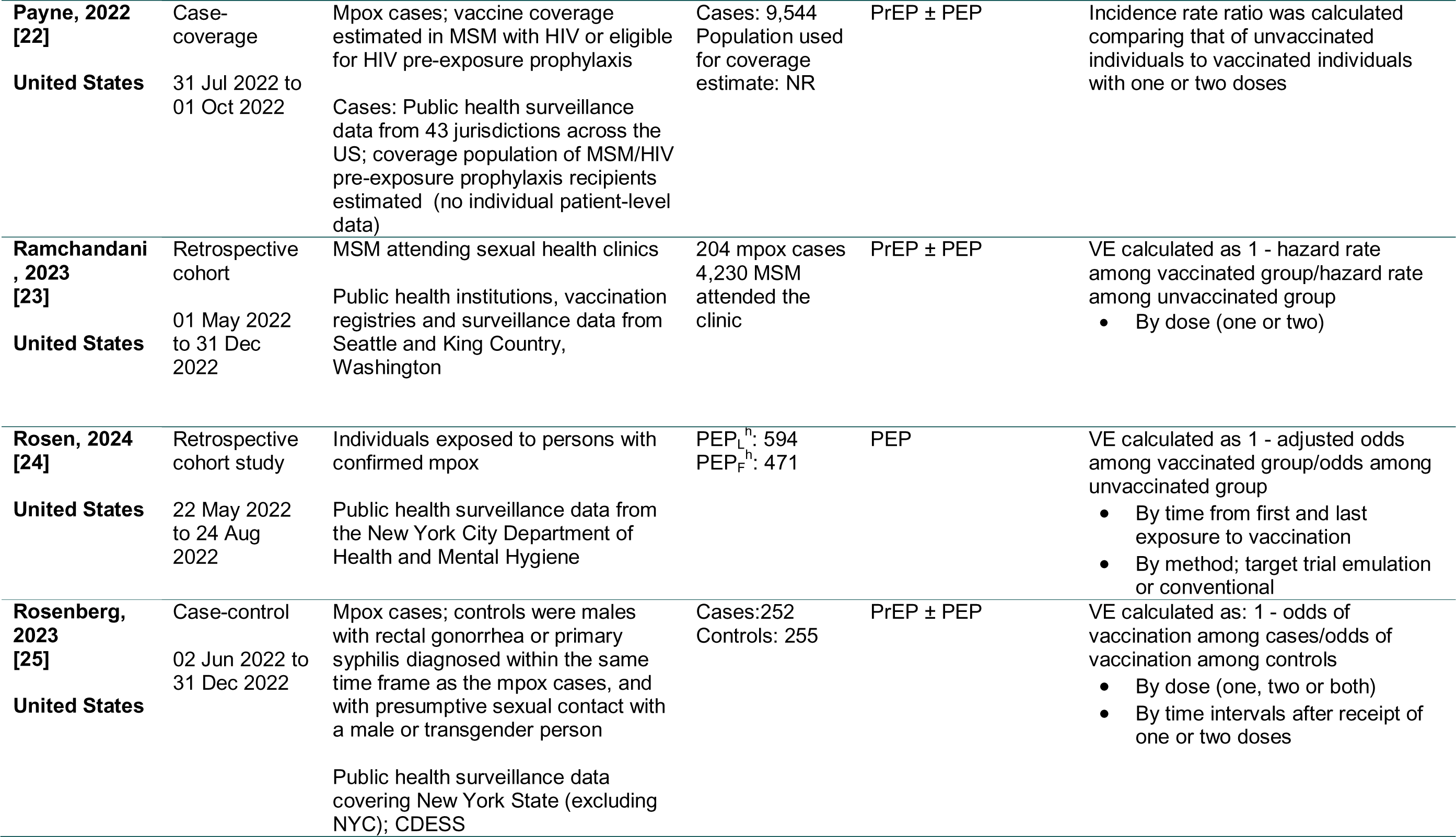
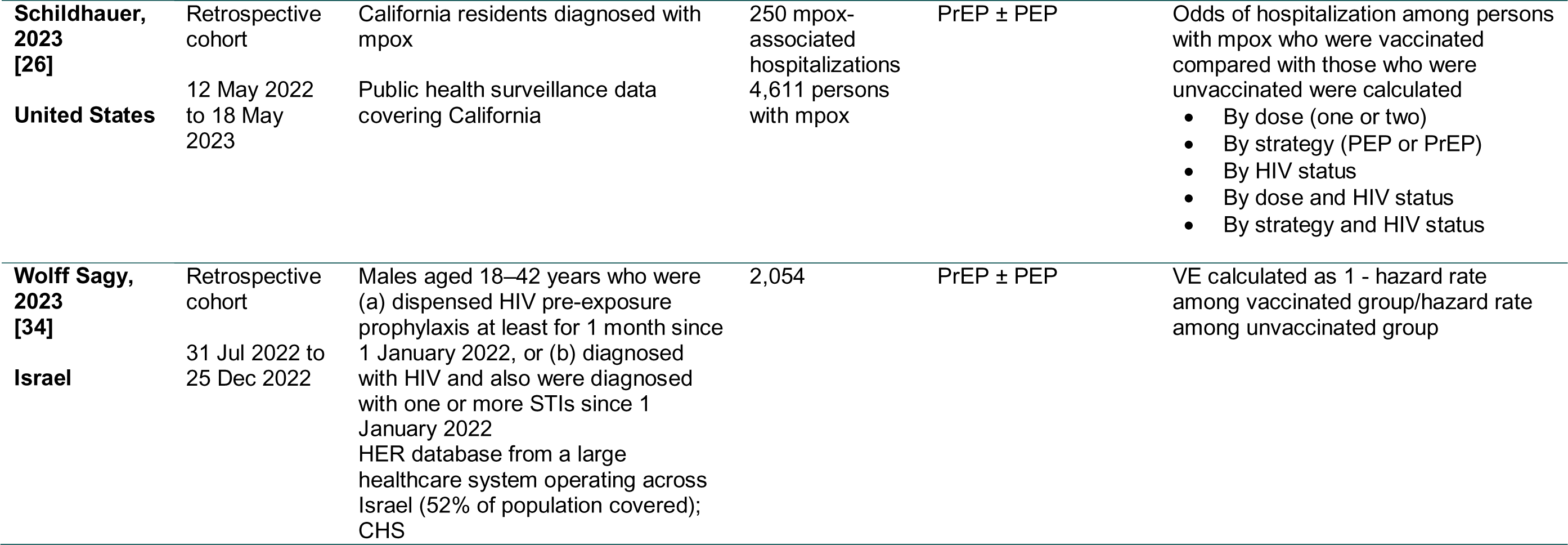
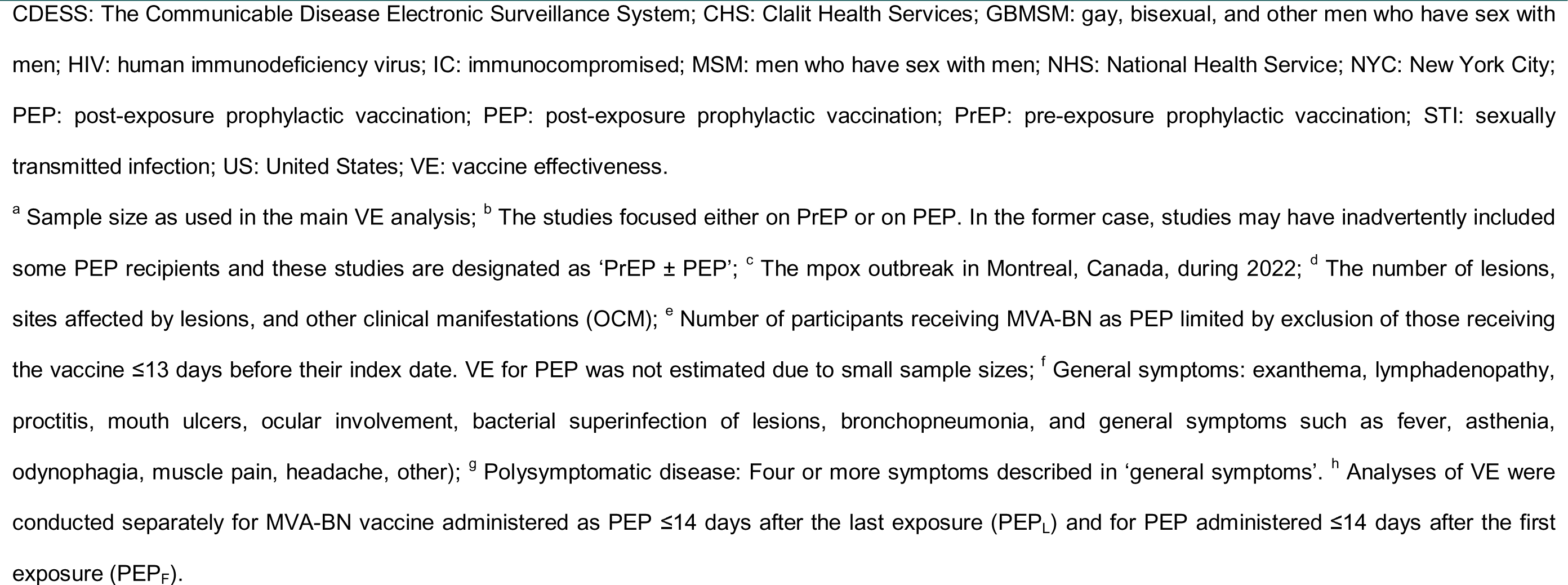
Characteristics of the studies included.

### Risk of bias

The risk of bias assessment found sources of (residual) confounding in several studies, with the most important ones resulting from the overall population setting and/or the selection of controls/comparator groups [21, 23–28, 31–34]. Sources of bias inherent to study design were also identified, including misclassification bias for studies using data from registries and databases [24, 25, 33], recall bias and non-response bias for studies using surveys and self-reported data [20, 24, 27, 28, 32], and an inability to control for confounders in case-coverage studies [22, 32, 33, 35]. Additionally, one study was found to be at risk of immortal time bias [31].

### Study results

#### Vaccine effectiveness estimates against symptomatic mpox infection for PrEP with MVA-BN

Twelve studies reported that PrEP with MVA-BN reduced the risk of symptomatic mpox infection at ≥14 days after vaccination. Half of the studies reported data for PrEP recipients only, while the other half may have included a proportion of PEP recipients (PrEP ± PEP). Five studies reported data for one dose of MVA-BN only, one study reported data for two doses only, and six studies reported data for both one and two doses (**Table 2**).

**Table 2.**
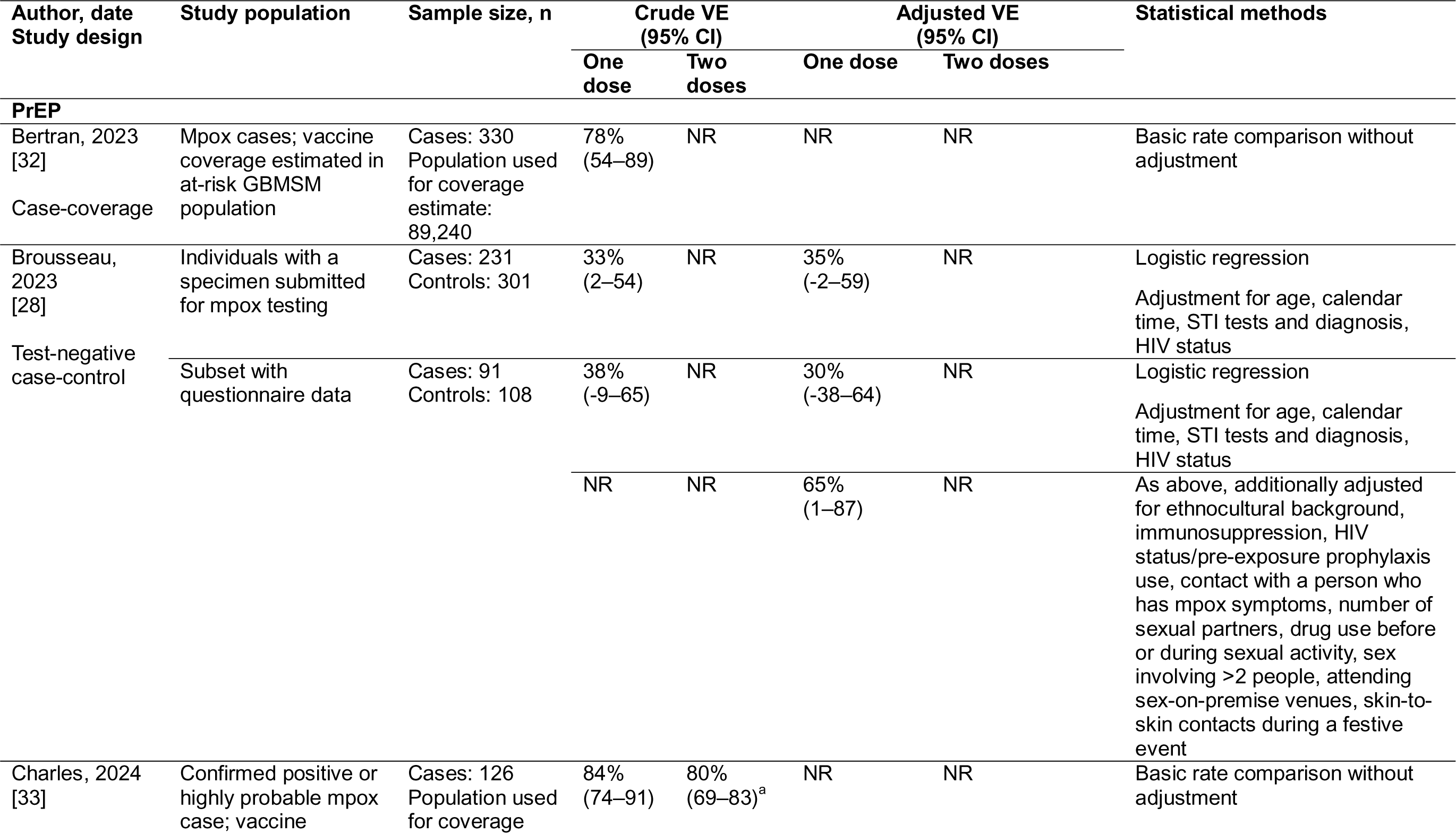
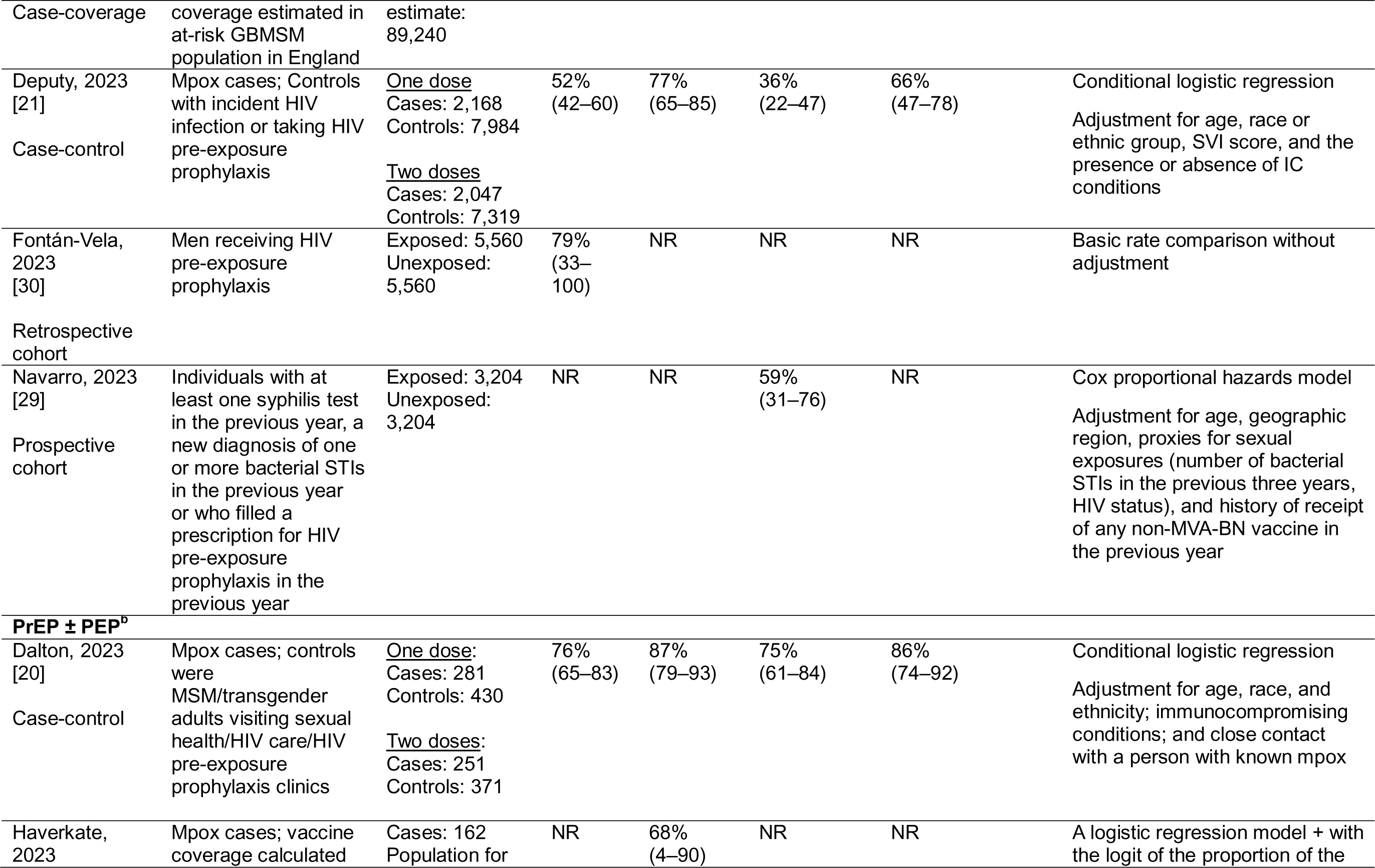
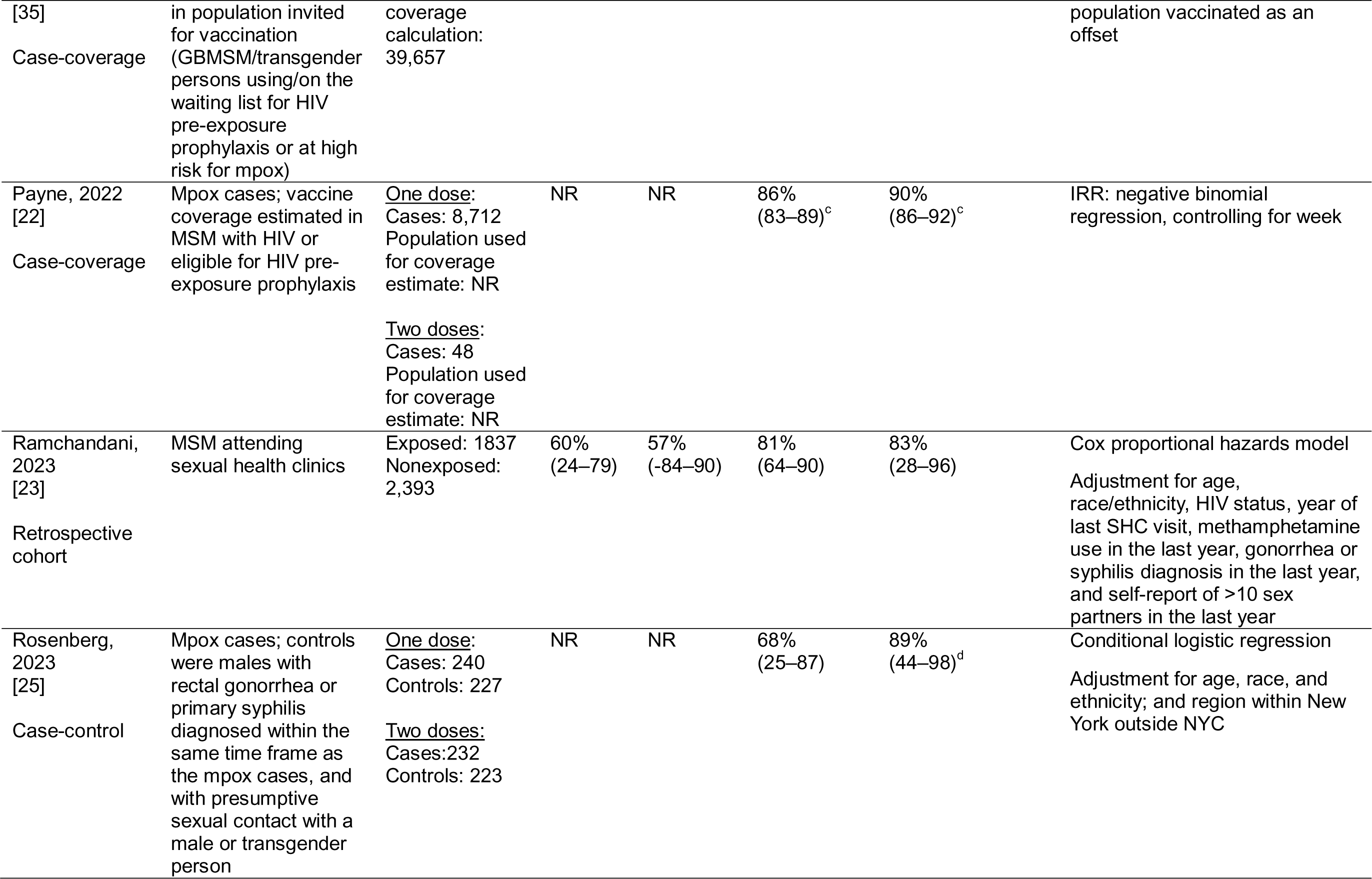
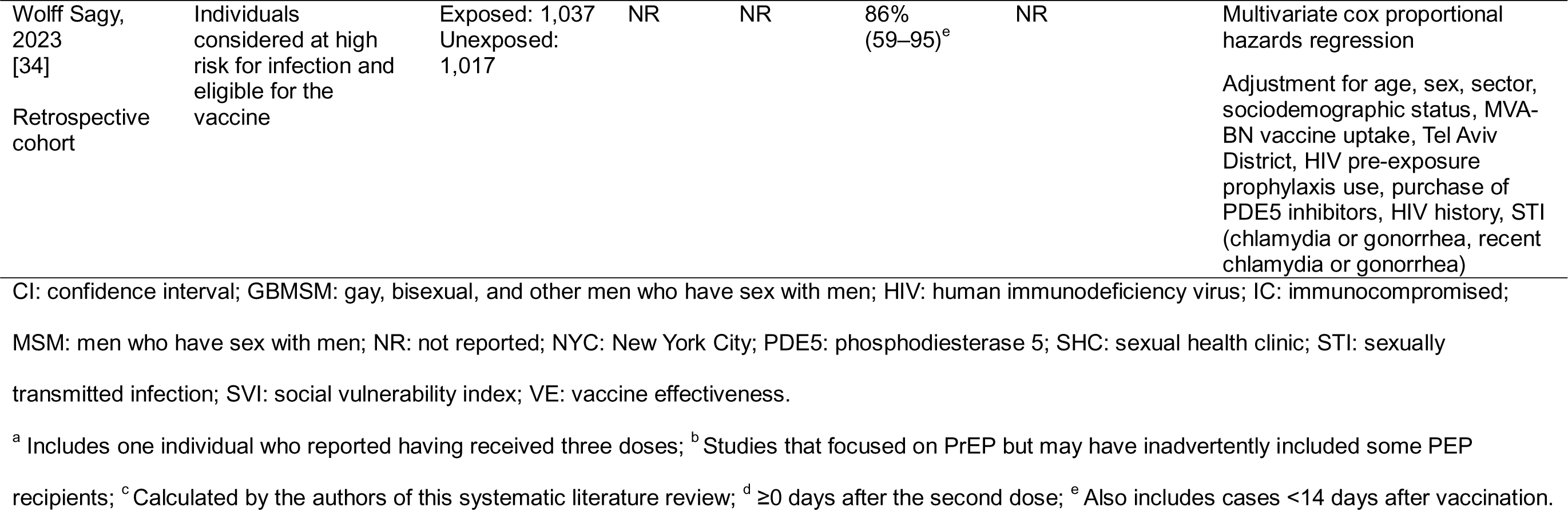
Vaccine effectiveness estimates against symptomatic mpox disease ≥ 14 days after vaccination for MVA-BN administered exclusively or primarily as pre-exposure prophylactic vaccination.

For one dose of MVA-BN administered as PrEP, adjusted VE estimates between 35% and 86% were reported across eight studies. Seven studies reported crude VE estimates, which fell within the same range (33%–84%). The studies conducted by Brousseau et al. [28] and Deputy et al. [21] reported adjusted VE estimates for one dose of MVA-BN of 35% and 36%, respectively, which were notably lower than the adjusted VE estimates reported in the other studies (≥59%) (**Table 2**).

For two doses of MVA-BN administered as PrEP, five studies reported adjusted VE estimates ranging from 66% to 90%. The reported crude VE estimates ranged from 57% to 87% in five studies (**Table 2**). The study by Charles et al. [33] was the only study to use data collected in the year 2023 (Jan 2023 until Dec 2023) only, and found crude VE estimates of 84% for one dose and 80% for two doses of MVA-BN (adjusted VE estimates were not reported) (**Table 2**).

#### Vaccine effectiveness estimates against symptomatic mpox infection for PEP with MVA-BN

Two studies reported on the effectiveness of one dose of PEP with MVA-BN against symptomatic mpox infection at ≥14 days after vaccination; no data were reported for two doses of PEP (**Table 3**).

**Table 3.** Vaccine effectiveness estimates against symptomatic mpox disease ≥14 days after second dose for MVA-BN administered as t-exposure prophylactic vaccination.

| Author, date<br>Study design | Study population | Sample size, n | Crude VE %<br>(95% CI) |  | Adjusted VE<br>(95% CI) |  | Statistical methods |
| --- | --- | --- | --- | --- | --- | --- | --- |
|  |  |  | One dose | Two doses | One dose | Two doses |  |
| Montero<br>Morales, 2023<br>[31]<br>Prospective<br>cohort study | Close contacts of<br>laboratory-confirmed mpox<br>cases | Exposed: 230<br>Unexposed: 254 | 84%<br>(66–92) | NR | 89%<br>(76–95)<sup>a</sup> | NR | Survival analysis<br>Adjustment for age, sex, type of close<br>contact, HIV pre-exposure<br>prophylaxis user, HIV infection, and<br>symptoms |
| Rosen, 2024<br>[24]<br>Retrospective<br>cohort study –<br>with target trial<br>design | Individuals having<br>contact with<br>persons with<br>confirmed mpox | PEP<sub>L</sub><sup>b</sup><br>Exposed: 333<br>Unexposed: 261<br>PEP<sub>F</sub><sup>b</sup><br>Exposed: 183<br>Unexposed: 288 | NR | NR | Conventional:<br>PEP<sub>L</sub>: 78%<br>(50–91)<br>PEP<sub>F</sub>: 73%<br>(31–91)<br>Target trial<br>PEP<sub>L</sub>: 19%<br>(-54–57)<br>PEP<sub>F</sub>: -7%<br>(-144–53) | NR | Conventional: multivariate logistic<br>regression, controlling for exposure<br>risk and race/ethnicity<br>Target trial emulation: sequence of<br>nested trials starting each day after<br>exposure to account for immortal time<br>bias. Logistic regression controlling<br>for exposure risk and race/ethnicity |

CI: confidence interval; HIV: human immunodeficiency virus; NR: not reported; PEP: post-exposure prophylaxis; VE: vaccine effectiveness.
<sup>a</sup> Also includes cases <14 days after vaccination; <sup>b</sup> Analyses of VE were conducted separately for MVA-BN vaccine administered as PEP $\leq 14$ days after the last exposure (PEP<sub>L</sub>) and for PEP administered $\leq 14$ days after the first exposure (PEP<sub>F</sub>).

The two studies found adjusted VE estimates of 78% and 89% when ‘classic statistical methods’ (multivariate logistic regression and survival analysis, respectively) were used. Rosen et al. [24] also calculated VE estimates using target trial emulation to account for immortal time bias, which were –7% for PEP administered ≤14 days after first exposure and 19% for PEP administered ≤14 days after last exposure; these VE estimates were associated with wide CIs due to small sample sizes (**Table 3**).

#### Vaccine effectiveness estimates of MVA-BN against mpox-related hospitalization and severity of mpox clinical manifestations

The study by Schildhauer et al. [26] reported a lower risk of mpox-related hospitalization among those who had been vaccinated with MVA-BN compared to those who had not been vaccinated. Based on ORs provided by Schildhauer et al., we calculated crude VE estimates against hospitalization of 73% and 80% for a vaccination schedule consisting of one or two MVA-BN doses, respectively, and 58% for one dose of MVA-BN administered as PEP <14 days before the episode date (**Table 4**).

**Table 4.**
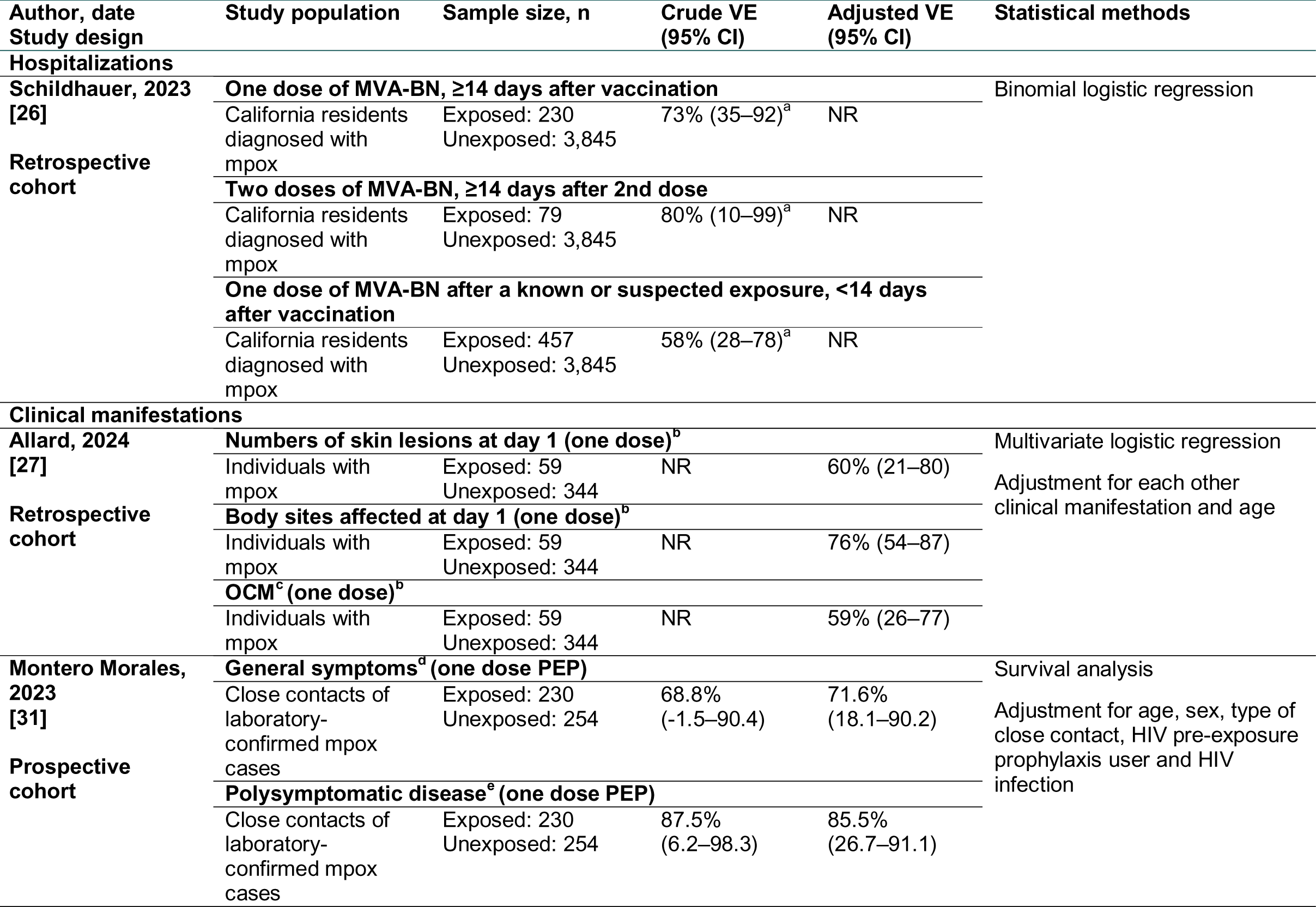
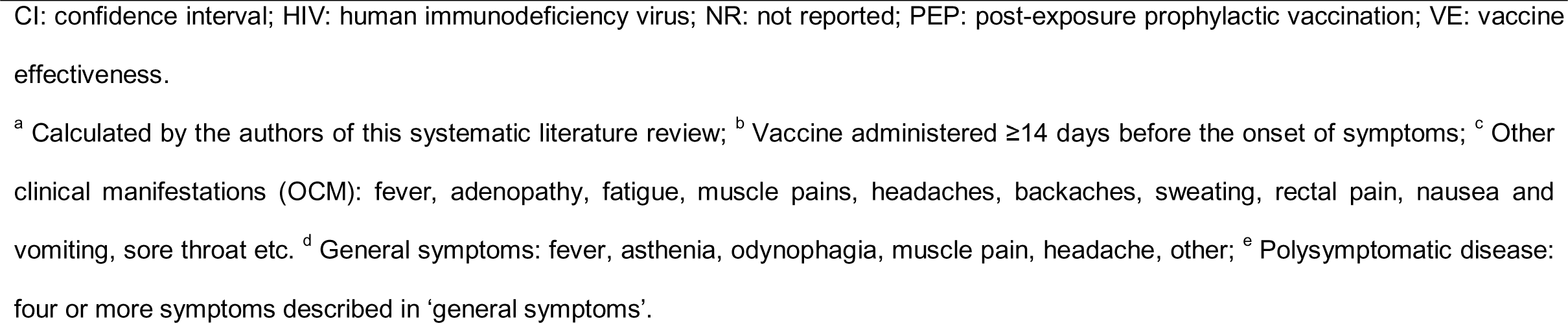
Vaccine effectiveness estimates against hospitalizations and mpox clinical manifestations.

Two papers reported a decreased severity of mpox clinical manifestations in cases who had received one dose of MVA-BN. Allard et al. [27] found adjusted VE estimates at day 1 of infection of 60% against the number of skin lesions, 76% against the number of body sites affected by skin lesions, and 59% against other clinical manifestations (incl. fever, muscle pain, and headache). Montero Morales et al. [31] reported adjusted VE estimates of one dose of MVA-BN administered as PEP of 72% against general symptoms (fever, asthenia, odynophagia, muscle pain, headache, or other symptoms) and 86% against polysymptomatic disease (i.e., occurrence of four or more general symptoms) (**Table 4**).

#### Vaccine effectiveness estimates of MVA-BN against symptomatic mpox infection by route of administration

Two studies reported data stratified by route of administration; one reported VE estimates while the other study only reported the number of mpox cases in vaccinated and unvaccinated groups (**Supplementary Table 1**). When adjusted VE estimates against symptomatic mpox infection were reported, these were in similar ranges for two doses of MVA-BN regardless of the route of administration: 80% (95% CI 23–95) for intradermal administration, 89% (95% CI 56–97) for subcutaneous administration, and 87% (95% CI 69–95) when administered via a combination of both routes (**Supplementary Table 1**).

#### Vaccine effectiveness estimates of MVA-BN against symptomatic mpox infection <14 days after vaccination

Four studies reported VE estimates stratified by time after vaccination, and as such demonstrated the risk of breakthrough infections before the onset of vaccine-induced immunity (**Supplementary Table 2**). Within the first 13 days post-vaccination, reported VE estimates for MVA-BN PrEP or ‘PrEP ± PEP’ did not exceed –4% (95% CI –50–29) (**Supplementary Table 2**).

## Discussion

This SLR identified 16 records on real-world studies assessing the VE of one or two doses of MVA-BN against symptomatic mpox infection, mpox-related hospitalization, and severity of mpox clinical manifestations. When MVA-BN was administered as PrEP, the adjusted VE estimates against symptomatic mpox infection ≥14 days post-vaccination were ≥35% (n=8 studies) for one dose and ≥66% (n=5) for two doses. The lower limit of the VE range for one dose of MVA-BN administered as PrEP was lower than that of other ranges because two studies reported VE estimates that were notably lower than other estimates [21, 28]. Outside of these data points, the adjusted VE estimates for PrEP with one dose of MVA-BN were ≥59%. Adjusted VE estimates against symptomatic mpox infection for MVA-BN administered as PEP were only reported for one dose and were ≥78% (n=2 studies). The VE estimates for two doses of MVA-BN fell in a similar range as VE estimates historically reported for replicating vaccinia-based smallpox vaccines [36]. Furthermore, MVA-BN vaccination reduced the risk of mpox-related hospitalization and the severity of mpox clinical manifestations.

In most studies, controls or unvaccinated comparator groups were selected based on characteristics such as MSM or transgender status, recent diagnosis of HIV or other STIs, and/or use of HIV pre-exposure prophylaxis. However, there was notable heterogeneity across studies in the definition of the at-risk population, which might have influenced the VE estimates. In the case-control study by Deputy et al. [21] for example, which found an adjusted VE estimate against symptomatic mpox infection of 36% for one dose of MVA-BN administered as PrEP and was one of the notably low VE estimates, controls were identified from a large secondary database that may not have had as granular information on risk proxies as studies using individual patient-level data. The control population was not restricted to MSM, resulting in a broader at-risk population than in other studies, which may have led to the inclusion of a higher proportion of subjects who were not vaccine-eligible in the control group and to the underestimation of the VE. This concern was also highlighted in a letter to the editor related to this article [37, 38]. The authors of this letter to the editor highlighted that the vaccine uptake of one dose of MVA-BN among controls reported by Deputy et al. (14.5%) was lower than vaccine uptake data reported by e.g., the CDC. When they recalculated VE estimates against symptomatic mpox infection using a vaccine uptake of 45.5% (i.e., uptake reported by CDC for the period until October 1, 2022 [22]), the crude VE estimate increased to fall within a similar range as VE estimates reported by other studies [22, 32, 34]. A very different approach regarding the included at-risk population was taken in the case-control study by Rosenberg et al. [25], where controls were restricted to MSM who had rectal gonorrhea or primary syphilis diagnosed within the same time frame as the mpox cases. This approach was assumed to lead to a control population that exhibited risk behavior similar to the cases within the same time period. A sensitivity analysis within that study included secondary syphilis cases in the control group and found a VE estimate for one or two MVA-BN doses against symptomatic mpox infection that trended lower than the main VE estimate, albeit with overlapping CIs (64.8% [95% CI 26.7–83.1] and 75.7% [95% CI 48.5–88.5], respectively). This difference may reflect the inclusion of control patients with more remote risk behaviors or different clinical presentations [25]. As a last example, the test-negative case-control study by Brousseau et al. [28] found a VE estimate against symptomatic mpox infection for one MVA-BN dose of 35% based on data in administrative databases only, with adjustment for surrogate indicators of exposure risk available in these databases. However, when the authors adjusted for self-reported risk factors, which were higher in number and more detailed than those available in the administrative databases, the VE estimate increased to 65% (with a wide CI) [28]. Though the sample size in this study was small, particularly for the sub-analysis (n=199 cases), the study design highlights the potential impact of confounding based on differential risk exposure by vaccination status.

Licensure of MVA-BN for protection against mpox was based on the efficacy of two MVA-BN doses against mpox observed in animal studies [39] and clinical safety and immunogenicity data. The latter includes the demonstration of non-inferior immunogenicity compared to the replicating-vaccinia vaccine ACAM2000 together with a more favorable safety profile for MVA-BN compared to ACAM2000 as demonstrated in a pivotal phase 3 randomized clinical trial [40]. While one dose of MVA-BN was previously shown to induce low titers of vaccinia-specific and MPVX-specific neutralizing antibody titers [41–43], our SLR found VE estimates against symptomatic infections for one dose of MVA-BN suggestive of high effectiveness. This challenges the view that neutralizing antibody levels correlate with protection against *Orthopoxvirus* infection. Indeed, previous experiments showed that MVA vaccination fully protected B cell-deficient mice in *Orthopoxvirus* challenge models, indicating that antibodies are not the sole correlate of protection [44]. Furthermore, a study including a small number of MVA-BN vaccinated individuals revealed that the protective immunogenicity of MVA-BN might be mostly mediated by T cells [45]. While a correlate of immunity for MVA-BN has not been established yet, these findings suggest that cellular immunity could be relevant. However, a pre-clinical animal study for an mRNA mpox vaccine candidate found that protection was primarily conferred by inducing a focused humoral immune response [46], which suggests that the correlate of immunity might depend on the vaccine platform.

This SLR employed a thorough process searching three literature databases combined with grey literature sources including preprints and a search through bibliographies of selected papers. Nevertheless, the presented data need to be interpreted in light of the high heterogeneity across included studies in terms of study design, characteristics of study populations, and study settings, among others, which limits comparisons. Because we considered that this heterogeneity across studies would hamper the interpretation of VE point estimates originating from MAs, we chose not to perform such analysis. This SLR also highlighted gaps in data. Most participants included in the identified studies were adult MSM, and data on females and children are lacking. The use of very specific at-risk groups reduced the generalizability of some studies. All identified studies took place in high-income countries, while no data from mpox-endemic countries were identified. Furthermore, several research groups initiated prospective studies in high-income countries in the early phases of the mpox outbreak, but these have not (yet) been finalized and/or published, primarily because of a waning number of cases. Most of these studies have now been suspended due to a lack of cases. Lastly, while we identified studies that demonstrated the risk of breakthrough infections within 14 days of vaccination, data on VE durability in real-world settings was limited. Only the study by Charles et al. from the UK provided some insight by reporting crude VE estimates against symptomatic infections of 84% and 80% for one and two doses of MVA-BN, respectively, based on confirmed and highly probable mpox diagnoses reported until December 2023, while most vaccines had been administered by March 2023 [33]. It should, however, be noted that follow-up time might impact VE estimates as a large proportion of those vaccinated will receive the vaccine during the early stages of its availability during an outbreak. Also, a larger proportion of those at greatest risk of infection are also likely to be infected earlier in the outbreak when awareness and vaccination coverage are lower. Therefore, a longer duration of follow-up may bias toward vaccine effect as overall case rates decline over time.

The included studies themselves also had limitations. A common limitation was that, due to the timing of the vaccination program within the mpox outbreak, limited numbers of cases were available for inclusion, especially after deployment of the second MVA-BN dose. This limited the studies in terms of power and the types of analyses that they could perform. However, all of the included studies were adequately powered for primary VE analyses. Furthermore, the studies were inconsistent in how they defined the use of MVA-BN as PEP and whether studies that focused on PrEP included or excluded PEP use in their analyses. The inclusion of a proportion of PEP recipients was unavoidable in some studies because of the data used for the analyses. In Germany, only 7% of all MVA-BN administrations from June 2022 to January 2024 were PEP [47]. Because the studies included in our SLR used mpox vaccination recommendations similar to those in place in Germany, it is likely that the majority of vaccine recipients included in ‘PrEP ± PEP’ studies received MVA-BN as PrEP. Lastly, all studies were found to be at risk of some (residual) confounding and other sources of bias. Many studies relied on data from databases, and exposure risk was defined based on proxies such as MSM or transgender status, recent diagnosis of HIV or other STIs, and/or use of HIV pre-exposure prophylaxis because individual-level behavior data were not available; however, this limited the researchers’ ability to account for confounding. Health-seeking behavior, HIV status, geographic location, socioeconomic status, race and gender identity are examples of other confounders which were not considered or accounted for by some studies. Studies that actively recruited participants could obtain more detailed data on exposure risk but were also subject to bias based on factors related to willingness to participate as well as recall bias. A test-negative case-control design, which might help reduce selection bias and avoid confounding associated with health-seeking behavior because cases and controls arise from the same source population, was employed in only one study [28]. Concern was raised in a letter to the editor that immortal time bias and confounding might have overestimated the adjusted VE of one dose of MVA-BN administered as PEP estimated by Montero Morales et al. (89% [95% CI 76–95]) [31, 48]. In their response letter, Montero Morales et al. performed an intent-to-treat analysis and found a VE of 75% (95% CI 55–86). Additionally, they performed a landmark approach analysis to try and account for immortal time bias by including vaccination as a time-varying exposure, which resulted in an estimated VE of 83% (95% CI 61–92) [49]. The importance of being mindful of immortal time bias was also demonstrated by the target trial emulation conducted by Rosen et al. [24], which lowered their adjusted VE estimates against symptomatic infections for PEP administered ≤14 days after last exposure from 78% (95% CI 50–91) to 19% (95% CI –54–57). Regardless of these limitations and the heterogeneity in study design and methods across the included studies, the VE estimates of MVA-BN against symptomatic mpox infection were generally high and in similar ranges. Taken together, the available real-world VE data support the use of MVA-BN in managing active outbreaks.

Future VE studies may be conducted if new outbreaks continue to expand. Ideally, investigators would prioritize maximizing comparability in risk of exposure/infection over time in the vaccinated and unvaccinated and in cases and controls in order to generate a more precise estimate of the true effectiveness of MVA-BN against mpox.

## Conclusions

Despite heterogeneity in study design, study settings, and at-risk populations across the 16 included studies, this SLR demonstrated the effectiveness of one or two doses of MVA-BN against symptomatic mpox infection in outbreak settings. As such, the presented data support the deployment of MVA-BN for mpox outbreak control. The identified methodological limitations and biases underscore the need for improved study designs to allow more accurate evaluation and more precise estimation of the true effectiveness of MVA-BN against mpox.

## Data Availability

All data used in this study originated from published sources.

## Additional information

## Abbreviations

CDC: US Centers for Disease Control and Prevention
CDESS: The Communicable Disease Electronic Surveillance System
CHS: Clalit Health Services
CI: confidence interval
DE: data extraction
DRC: Democratic Republic of Congo
FT: full-text review
GBMSM: gay, bisexual, and other men who have sex with men
HIV: human immunodeficiency virus
HRR: hazard rate ratio
IC: immunocompromised
IRR: incidence rate ratio
MeSH: medical subject headings
MPVX: monkeypox virus
MSM: men who have sex with men
MVA-BN: Modified Vaccinia Ankara-Bavarian Nordic
NR: not reported
NYC: New York City
OCM: other clinical manifestations
OR: odds ratio
PDE5: phosphodiesterase 5
PEP: post-exposure prophylactic vaccination
PrEP: pre-exposure prophylactic vaccination
PICOS: patient, intervention, comparison, outcome, setting
PRISMA: Preferred Reporting Items for Systematic Reviews and Meta-Analyses
RR: risk ratio
RWE: real-world evidence
SHC: sexual health clinic
SLR: systematic literature review
STI: sexually transmittable infection
SVI: social vulnerability index
VE: vaccine effectiveness

## Acknowledgments

The authors thank Lotte Mathé (P95) for medical writing support.

## Author contributions

FL: Conceptualization; Writing – review and editing

SS: Conceptualization; Writing – review and editing

EB: Investigation; Writing – original draft; Writing – review and editing

LM: Investigation; Writing – original draft; Writing – review and editing

MRM: Conceptualization; Supervision; Writing – review and editing

## Availability of data

Other: All data used in this study originated from published sources.

