## Supplementary Methods for "MVA-BN Vaccine Effectiveness: A Systematic Review of Real-World Evidence in Outbreak Settings"

#### Full search strategy per database

| **PubMed** | | | | |
| --- | --- | --- | --- | --- |
| Date of last search: 28 Feb 2024 | | | | |
| **Search number** | | **Query** | | **Hits** |
| #4 | | #3 AND Filters: from 2022/01/01 to 2024/02/28 | | **546** |
| #3 | | #1 AND #2 | | 775775 |
| #2 | | "vaccine effectiveness" OR VE OR (effect* AND (vaccines[Mesh] OR vaccin* OR immuniz* OR immunis*)) | | 1,013,376 |
| #1 | | monkeypox OR monkeypox[Mesh] OR mpox | | 4,549 |
| **EMBASE** | | | | |
| Date of last search: 28 Feb 2024 | | | | |
| **Search number**  **^O^** | **Query** | | **Hits** | |
| #4 | #3 AND []/01-01-2022sd  NOT [29-02-2024]/sd | | **614** | |
| #3 | #1 AND #2 | | 713 | |
| #2 | 'vaccine effectiveness'/exp OR 'vaccine effectiveness' OR ve OR (('vaccine'/exp OR vaccin* OR immunis* OR immuniz*) AND effect*) | | 403,017 | |
| #1 | 'monkeypox'/exp OR monkeypox OR mpox | | 5,494 | |
| **LILACS** | | | | |
| Date of last search: 28 Feb 2024 | | | | |
| **Search number**  **^O^** | **Query** | | **Hits** | |
| #3 | 2022–2024 | | **13** | |
| #2 | #1 AND Limits "LILACS" | | 17 | |
| #1 | ((monkeypox OR mpox OR "monkeypox virus") AND ("vaccine effectiveness" OR vaccine OR effectiveness)) | | 1,089 | |

#### Eligibility criteria

| **PICOS** | **Inclusion criteria** | **Exclusion criteria** |
| --- | --- | --- |
| Population | Vaccinated and unvaccinated population (non-restricted) | None |
| Intervention | Jynneos®/Imvanex®/Imvamune® vaccination | Other mpox vaccines |
| Comparator | Non-vaccinated by Jynneos®/Imvanex®/Imvamune®, vaccine | Studies looking at only vaccinated or only unvaccinated populations, e.g., case series |
| Outcomes | Vaccine effectiveness (VE)^a^ | Studies not reporting the outcome of interest |
| Study design | Observational studies: Case-control studies, prospective and retrospective cohort studies, and cross-sectional studies | Peer-reviewed publications that do not clearly outline methods and sources for data collection/analysis |
|  |  | - *In vitro* or modeling studies - SLRs^b^, randomized and non-randomized clinical trials - News and opinion articles - Case reports - Case series - Narrative reviews, letters |
| **Other criteria** | | |
| Publication period | Jan 1, 2022 – Feb 28, 2024 | Study period before 2022 |
| Language | English | Other languages |
|  | Spanish |  |
|  | French |  |
| Countries | Non-restricted | Not applicable |

^a^ Against clinical disease, not serological results; ^b^ Systematic literature reviews covering the objective were included during the title and abstract screening phase to identify additional relevant studies within the reference lists

#### Grey literature search

| Website/source | Search terms and limits | Screening approach | Relevant articles + not included in literature search |
| --- | --- | --- | --- |
| **Grey literature** | | | |
| Open Grey.UK^a^ | (monkeypox OR mpox) AND (vaccine AND effectiveness) | All hits were screened, filtered by year | No relevant additional articles found |
| ECDC website^a^ | (monkeypox OR mpox) AND (vaccine AND effectiveness) | All hits were screened, filtered by last year and by latest | No relevant additional articles found |
| CDC website^a^ | NA | Website, specifically the MVA-BN vaccine effectiveness page, was checked for relevant publications | No relevant additional articles found |
| Google scholar^a^ | (monkeypox OR mpox) AND (vaccine AND effectiveness), | First five pages of results were screened | **One relevant record retained** |
| MedRxiv & BioRxiv^a^ | (monkeypox OR mpox) AND (vaccine AND effectiveness) | All hits were  screened and filtered  by newest | **Four relevant records retained** |
| MMWR CDC^a^ | NA | Latest volumes were checked | No relevant additional articles found |
| **Hand search – from reference lists of included articles** | | | |
| NA | NA | NA | **One relevant record retained** |
| **Provided by experts** | | | |
| NA | NA | NA | **Three relevant records retained** |
| CDC: Centers for Disease Control and Prevention; ECDC: European Centre for Disease Control and Prevention; MMWR: The Morbidity and Mortality Weekly Report; NA: not applicable.  ^a^ date of last search: March 5, 2024 | | | |
