## Supplementary Table 1 for "MVA-BN Vaccine Effectiveness: A Systematic Review of Real-World Evidence in Outbreak Settings"

### Supplementary table 1. Vaccine effectiveness estimates against symptomatic mpox infection of MVA-BN by route of administration

| Author, date  Study design | Study population | Sample size, n | Crude VE (95% CI) | Adjusted VE (95% CI) | Statistical methods |
| --- | --- | --- | --- | --- | --- |
| Subcutaneous administration | | | | | |
| Deputy, 2023  [1]  Case-control | Mpox cases; controls with incident HIV infection or taking HIV pre-exposure prophylaxis | Cases: 2,028  Controls: 7,047 | 1 or 2 doses: NR^a^ | 1 or 2 doses: NR^a^ | Basic rate comparison without adjustment |
| Dalton, 2023  [2]  Case-control | Mpox cases; controls were MSM/transgender adults visiting sexual health/HIV care/HIV pre-exposure prophylaxis clinics | Cases: 268  Controls: 379 | 1 dose: 76%  (61–85)  2 doses: 89%  (61–97) | 1 dose: 77% (60–87)  2 doses: 89% (56–97) | Conditional logistic regression  Adjustment for age, race and ethnicity, immunocompromising conditions, and close contact with a person with known mpox |
| Intradermal administration | | | | | |
| Deputy, 2023  [1]  Case-control | Mpox cases; Controls with incident HIV infection or taking HIV pre-exposure prophylaxis | Cases: 2,227  Controls: 7,026 | 1 or 2 doses: NR^a^ | 1 or 2 doses: NR^a^ | Basic rate comparison without adjustment |
| Dalton, 2023  [2]  Case-control | Mpox cases; controls were MSM/transgender adults visiting sexual health/HIV care/HIV pre-exposure prophylaxis clinics | Cases: 246  Controls: 294 | 1 dose: 77%  (57–88)  2 doses: 81%  (38–94) | 1 dose: 81% (56–91)  2 doses: 80% (23–95) | Conditional logistic regression  Adjustment for age, race and ethnicity, immunocompromising conditions, and close contact with a person with known mpox |
| Heterologous administration | | | | | |
| Deputy, 2023  [1]  Case-control | Mpox cases; Controls with incident HIV infection or taking HIV pre-exposure prophylaxis | Cases: 2,030  Controls: 7,134 | 2 doses: 84%  (67–92) | 2 doses: 75% (48–88) | Conditional logistic regression  Adjustment for age, race or ethnic group, SVI score, and the presence or absence of IC conditions |
| Dalton, 2023  [2]  Case-control | Mpox cases; controls were MSM/transgender adults visiting sexual health/HIV care/HIV pre-exposure prophylaxis clinics | Cases: 239  Controls: 318 | 2 doses: 88%  (76–94) | 2 doses:87% (69–95) | Conditional logistic regression  Adjustment for age, race and ethnicity, immunocompromising conditions, and close contact with a person with known mpox |
| CI: confidence interval; HIV: human immunodeficiency virus; IC: immunocompromised; MSM: men who have sex with men; NR: not reported; SVI: social vulnerability index; VE: vaccine effectiveness.  ^a^ No. of mpox cases were reported for unvaccinated and vaccinated groups but VE estimates were not reported. | | | | | |

1. Deputy, N.P., et al., *Vaccine Effectiveness of JYNNEOS against Mpox Disease in the United States.* N Engl J Med, 2023. **388**(26): p. 2434-2443.
2. Dalton, A.F., et al., *Estimated Effectiveness of JYNNEOS Vaccine in Preventing Mpox: A Multijurisdictional Case-Control Study - United States, August 19, 2022-March 31, 2023.* MMWR Morb Mortal Wkly Rep, 2023. **72**(20): p. 553-558.
