## Supplementary Table 2 for "MVA-BN Vaccine Effectiveness: A Systematic Review of Real-World Evidence in Outbreak Settings"

### Supplementary table 2. Vaccine effectiveness estimates against symptomatic mpox infection of MVA-BN by time after vaccination

| Author, date  Study design | Study population | Sample size, n | Time stratification | Crude VE (95% CI) | Adjusted VE (95% CI) |
| --- | --- | --- | --- | --- | --- |
| Fontán-Vela, 2023  [1]  Retrospective cohort | Men receiving HIV pre-exposure prophylaxis | Exposed: 5,560  Unexposed: 5,560 | 0–6 days after 1^st^ dose | -38%  (-333–46) | NR |
|  |  | Exposed: 5,560  Unexposed: 5,560 | 0–13 days after 1^st^ dose | -14%  (-200–48) | NR |
|  |  | Exposed: 5,560  Unexposed: 5,560 | ≥7 days after 1^st^ dose | 65%  (23–88) | NR |
|  |  | Exposed: 5,560  Unexposed: 5,560 | ≥14 days after 1^st^ dose | 79%  (33–100) | NR |
| Navarro, 2023  [2]  Prospective cohort | Individuals with at least one syphilis test in the previous year, a new diagnosis of one or more bacterial STIs in the previous year or who filled a prescription for HIV pre-exposure prophylaxis in the previous year | Exposed: 3,204  Unexposed: 3,204 | 0–14 days after 1^st^ dose | NR | -25% (-114–27) |
|  |  | Exposed: 3,204  Unexposed: 3,204 | >14 days after 1^st^ dose | NR | 59% (31–76) |
| Rosenberg, 2023  [3]  Case-control | Mpox cases; Controls were males with rectal gonorrhea or primary syphilis diagnosed within the same time frame as the mpox cases, and with presumptive sexual contact with a male or transgender person | Cases: 240  Controls: 213 | <14 days after 1^st^ dose | NR | -36% (<-100–56) |
|  |  | Cases: 240  Controls: 227 | ≥14 days after 1^st^ dose | NR | 68% (25–87) |
| Bertran, 2023  [4]  Case-coverage | Mpox cases; vaccine coverage estimated in at-risk GBMSM population | Cases: 354  Population used for coverage estimate: 89,240 | <14 days after 1^st^ dose | -4% (-50–29) | NR |
|  |  | Cases: 330  Population used for coverage estimate: 89,240 | ≥14 days after 1^st^ dose | 78% (54–89) | NR |
| CI: confidence interval; GBMSM: gay, bisexual, and other men who have sex with men; HIV: human immunodeficiency virus; NR: not reported; STI: sexually transmitted infection; VE: vaccine effectiveness. | | | | | |

1. Fontán-Vela, M., et al., *Effectiveness of Modified Vaccinia Ankara-Bavaria Nordic Vaccination in a Population at High Risk of Mpox: A Spanish Cohort Study.* Clin Infect Dis, 2024. **78**(2): p. 476-483.
2. Navarro, C., et al., *Effectiveness of one dose of MVA-BN vaccine against mpox infection in males in Ontario, Canada: A target trial emulation*. 2023.
3. Rosenberg, E.S., et al., *Effectiveness of JYNNEOS Vaccine Against Diagnosed Mpox Infection - New York, 2022.* MMWR Morb Mortal Wkly Rep, 2023. **72**(20): p. 559-563.
4. Bertran, M., et al., *Effectiveness of one dose of MVA-BN smallpox vaccine against mpox in England using the case-coverage method: an observational study.* Lancet Infect Dis, 2023. **23**(7): p. 828-835.
